# Erythrocyte n-6 polyunsaturated fatty acids, gut microbiota and incident type 2 diabetes: a prospective cohort study

**DOI:** 10.1101/2020.03.29.20039693

**Authors:** Zelei Miao, Jie-sheng Lin, Yingying Mao, Geng-dong Chen, Fang-fang Zeng, Hong-li Dong, Zengliang Jiang, Jiali Wang, Congmei Xiao, Menglei Shuai, Wanglong Gou, Yuanqing Fu, Fumiaki Imamura, Yu-ming Chen, Ju-Sheng Zheng

**Affiliations:** School of Life Sciences, Westlake University, Hangzhou, China; Guangdong Provincial Key Laboratory of Food, Nutrition and Health; Department of Epidemiology, School of Public Health, Sun Yat-sen University, Guangzhou, China; Department of Epidemiology and Biostatistics, School of Public Health, Zhejiang Chinese Medical University, Hangzhou, China; Department of Epidemiology, School of Basic Medical Sciences, Jinan University, Guangzhou, China; Institute of Basic Medical Sciences, Westlake Institute for Advanced Study, Hangzhou, China; MRC Epidemiology Unit, University of Cambridge, Cambridge, UK

## Abstract

**Objective:** To examine the association of erythrocyte n-6 polyunsaturated fatty acid (PUFA) biomarkers with incident type 2 diabetes (T2D) and explore the potential role of gut microbiota in the association.

**Design:** We evaluated 2,731 non-T2D participants recruited between 2008-2013 in the Guangzhou Nutrition and Health Study, China. T2D cases were identified with clinical and biochemical information collected at follow-up visits. Using stool samples collected during the follow-up in the subset (n=1,591), 16S rRNA profiling was conducted. Using multivariable-adjusted Poisson or linear regression, we examined associations of erythrocyte n-6 PUFA biomarkers with incident T2D, and diversity and composition of gut microbiota.

**Results:** Over 6.2 years of follow-up, 276 T2D cases were identified (risk=0.10). Higher levels of erythrocyte γ-linolenic acid (GLA), but not linoleic or arachidonic acid, were associated with higher T2D incidence. Comparing the top to the bottom quartile groups of GLA levels, relative risk was 1.72 (95% confidence intervals: 1.21, 2.44) adjusted for potential confounders. Baseline GLA was inversely associated with gut microbial richness and diversity (α-diversity, both *p*<0.05) during follow-up, and significantly associated with microbiota β-diversity (*p*=0.002). Seven genera (*Butyrivibrio, Blautia, Oscillospira, Odoribacter, S24-7 other, Rikenellaceae other*, and *Clostridiales other*) were enriched in quartile 1 of GLA, and in participants without T2D.

**Conclusion:** Relative concentrations of erythrocyte GLA were positively associated with incident T2D in a Chinese population and also with gut microbial profiles. These results highlight that gut microbiota may play an important role linking n-6 PUFA metabolism and T2D etiology.

## INTRODUCTION

Type 2 diabetes (T2D) is one of the most prevalent metabolic conditions worldwide, and 463 million adults are living with diabetes. ^1^ To examine lifestyle-related risk factors for the development of T2D, the role of different fatty acids has been an important research agenda. During the past two decades, consumption of n-6 polyunsaturated fatty acids (PUFAs), rich in vegetable oil, has been increasing rapidly,^2 3^ while their relationship with human health remains controversial. The metabolic pathway of n-6 PUFAs is hypothesized to be closely involved in the T2D etiology,^4^ but has not been well characterized yet.

A recent pooled analysis of 20 prospective cohort studies investigated the association of linoleic acid ((LA; 18:2n6) and arachidonic acid (AA; 20:4n6) biomarkers with incident T2D, reporting that LA, but not AA, was inversely associated with T2D.^5^ However, in this pooling project, only two individual n-6 PUFA biomarkers were evaluated while cohorts in Europe reported gamma-linolenic acid (GLA) and other n-6 PUFAs to be associated with higher T2D risk.^5^ Moreover, with only one small cohort from Taiwan in the above pooling project,^5^ evidence from Asian populations has been limited. As an Asian population has different metabolic and lifestyle characteristics in relation to diabetes compared to American or European populations,^6^ more investigation for n-6 PUFA biomarkers in Asian populations is highly warranted.

The protective effect of n-6 PUFA or LA, the most abundant n-6 PUFA, on insulin homeostasis has been well characterized and discussed to link n-6 PUFA exposure to lower T2D risk.^7 8^ However, gut microbiota has been little investigated in research on PUFAs and any non-communicable diseases. On one hand, gut microbiota has been considered as an important risk factor for T2D and insulin resistance.^9 10^ On the other, rodent studies have suggested that high tissue levels of n-6 PUFAs are associated with differences in gut microbiota, including increased *Proteobacteria*, reduced *Actinobacteria* and *Bacteroidetes*, and altered microbial α-diversity.^11–13^ Yet, to our knowledge, there are no cohort studies investigating the potential link between n-6 PUFA exposure, T2D risk, and gut microbiota composition and diversity.

Evaluating the prospective cohort in China, we aimed to examine the prospective associations 1) between n-6 PUFA biomarkers and T2D risk in a Chinese population, and 2) between n-6 PUFA biomarkers and gut microbiota composition and diversity, and subsequent health outcomes.

## METHODS

### Study design and population

This study was based on the Guangzhou Nutrition and Health Study (GNHS), a community-based prospective cohort in southern China. Detailed information of the participants and study design was described previously.^14^ Briefly, 4,048 Chinese participants aged 40-75 years and lived in Guangzhou for at least 5 years were recruited between 2008-2010 (n=3,169) and 2012-2013 (n=879). All participants have been followed up every 3 years approximately. During each visit, demographic, dietary and lifestyle information was collected via questionnaires. The study was registered at clinicaltrials.gov (NCT03179657).

According to our pre-specified inclusion and exclusion criteria of our participants for the present analysis (Figure S1), we excluded participants with missing information on age or sex (n=9), diet (n=7), self-reported/diagnosed T2D (n=400) or history of cancer (n=16) at baseline, or extreme levels of self-reported total energy intake (<800 kcal or >4,000 kcal per day for men, and <500 kcal or > 3,500 kcal per day for women) (n=53). We also excluded those without measurement of erythrocyte membrane fatty acid compositions at baseline (n=298) and further excluded those without any follow-up information on T2D status (n=534, follow-up rate 84%). The current analyses were censored by date of T2D ascertainment or Apr 30, 2019, which ever happened first. A total of 2,731 individuals were included in the present study, with a median follow-up time of 6.2 years.

Among the above included 2,731 individuals, stool samples from 1,606 individuals were collected between 2014-2018 for the subsequent 16S profiling, with 1,591 individuals included in the microbial association analysis after excluding participants who used antibiotics within two weeks before stool collection (n=15).

### Measurement of erythrocyte membrane fatty acids and covariates

We considered relative concentrations (% of total fatty acids) of erythrocyte LA, GLA, and AA as the main exposure variables. Blood samples were drawn after overnight fasting at the baseline visit. Erythrocytes were aliquoted within 2 hours of blood sampling and stored at −80°C. As previously described,^15^ fatty acid moieties of erythrocyte membranes were trans-methylated and measured as proportions (%) of total fatty acids by using gas chromatography (7890 GC, DB-23 capillary column 60m×0.25mm internal diameter×0.15μm film, Agilent, California, USA). Commercially available standards (Nu-Chek Prep, Minnesota, USA) were used to identify individual fatty acids (n=37) and quantify a relative peak strength of each. The intra-assay coefficients of variation for LA, GLA and AA were 6.4%, 12.8%, and 8.04%, respectively. Sociodemographic, lifestyle, and clinical covariates were collected with standardized questionnaires. Details of fatty acid and covariate measurements are available in Supplementary Methods.

### T2D case ascertainment

At baseline (for exclusion) and follow-up visits, T2D cases were ascertained according to the criteria of American Diabetes Association for T2D diagnosis^16^ if a participant met one of the following criteria: a fasting blood glucose concentration ≥7.0 mmol/L (126 mg/dL), or HbA1c concentration ≥6.5%, or self-reported medical treatment for diabetes.

### Fecal sample collection and 16S rRNA profiling

Fecal samples were collected on site during follow-up visits between 2014-2018. Microbial DNA extraction, polymerase chain reaction (PCR) and amplicon sequencing were performed as described (see Supplementary Methods). Fastq-files were demultiplexed, merge-paired, quality filtered by Quantitative Insights into Microbial Ecology software (version 1.9.0).^17^ Sequences were clustered into operational taxonomic units (OTUs) with 97% similarity and annotated based on the Greengenes Database (version 13.8).^18^

### Statistical analysis

Statistical analyses were performed using Stata 15 (StataCorp, Texas, USA) or R (version 3.6.3). We used multivariable Poisson regression models to estimate the risk ratio (RR) and 95% confidence interval (CI) of T2D comparing quartiles of the erythrocyte n-6 PUFA (LA, GLA and AA), adjusting for potential confounders. We fitted three statistical models: model 1 included age, sex, BMI, and waist-hip ratio (WHR); model 2, model 1 + education, household income, smoking, alcohol drinking status, physical activity, total energy intake, and family history of diabetes; and model 3, as model 2 + baseline erythrocyte total n-3 PUFAs and fasting glucose. *p*-value for trend was calculated based on per quartile difference in the corresponding n-6 PUFA variable.

In sensitivity analyses, we repeated the above analysis based on model 3 to examine the impact of loss to follow up by a simple imputation (assuming that participants loss to follow up did not develop T2D), and 10 rounds of multiple imputations based on regression model using all covariates listed in model 3.

We assessed influences of additional potential covariates on the models: dietary variables (dietary intake of dairy products, red and processed meat, fish, vegetable, and fruit, in quartiles), blood lipids (total triglycerides and low-density lipoprotein), prevalent coronary heart disease, treatment for hypertension or hyperlipidemia. We further examined the potential interaction of erythrocyte n-6 PUFA biomarkers with age, sex, BMI or n-3 PUFA biomarker concentrations on T2D risk. Post-hoc stratified analyses were performed if there was a significant interaction (*p*_*interaction*_<0.05) for age, sex, BMI or n-3 PUFA biomarker concentrations with the n-6 PUFA biomarkers.

For the gut microbiome analysis, we calculated the α-diversity metrics (observed OTUs, Shannon’s diversity index, Chao index and Simpson index) by using QIIME (version 1.9.0) based on rarefied OTU counts. We standardized α-diversity indicators (Z-score was calculated: the variable was subtracted by the mean and divided by the standard deviation) and then reduced assay-dependent variabilities by fitting a linear mixed model with an α-diversity indicator as an outcome variable and with assay-specific covariates including sequencing depth and Bristol scale as fixed effects, and sequencing batch as a random effect. The residuals from this modeling were then used in the following n-6 PUFA biomarker association analyses. We used a linear regression model to examine the association of individual n-6 PUFA biomarkers (by quartile) with the four α-diversity metrics, adjusted for age, sex, BMI, waist-hip ratio, education, household income, smoking status, alcohol drinking status, physical activity, total energy intake, and baseline erythrocyte total n-3 PUFAs. We also conducted an additional adjustment for dietary fiber intake to assess the potential marginal effect of dietary fiber, as fiber is a well-known factor influencing the gut microbiome.^19^ To investigate the relevance of significant findings in relation to T2D, we performed mediation analysis (R {*mediation*})^20^ with the residuals of α-diversity as a mediator of the relationship between n-6 PUFAs and T2D, adjusting for all covariates described in model 3. We examined the cross-sectional associations between the residuals of α-diversity metrics and T2D by using logistic regression models, adjusted for age, sex, BMI, education, household income, smoking status, alcohol drinking status, prevalent hypertension and dyslipidemia.

We conducted a principal coordinate analysis (PCoA) and permutational multivariate analysis of variance (PERMANOVA) (R function adonis {*vegan*}, 999 permutations)^21^ based on Bray-Curtis distance to compare the whole gut composition (β-diversity) at genus level by quartiles of n-6 PUFA biomarkers. The potential confounders included in the PERMANOVA were sequencing depth, sequencing batch, Bristol scale, age, sex, BMI, WHR, education, household income, smoking status, alcohol drinking status, physical activity, total energy intake, and baseline erythrocyte total n-3 PUFAs. The above analysis was repeated for gut composition at OTU level. Scaled relative abundances (i.e., divided by the standard deviation) were used to lessen the influence of highly abundant genera on Bray-Curtis distance. The association of community structure with T2D was also assessed by PERMANOVA (R function adonis, 999 permutations) to obtain R^2^ indicating the proportion of the variability explained by the studied variables including n-6 PUFAs. We performed the post-hoc pairwise comparisons between different quartiles of GLA with Bonferroni correction (cut-off *p*<0.008).

At genus level, we performed a linear discrimination analysis (LDA) as implemented using LEfSe.^22^ The default parameters were used (α value for Wilcoxon tests was 0.05 and the LDA score was 2.0) to identify biomarkers at genus level distinguishing different quartiles of n-6 PUFAs. Only taxon present in at least 10% of samples were included in the analyses. Then, we compared the relative abundances of genera by T2D status using the LEfSe method.^22^ Besides, we evaluated the potential correlation of the above identified genera with T2D-related traits, including fasting insulin, glucose, total cholesterol, total triglycerides, low-density lipoprotein, high-density lipoprotein cholesterol (HDL), non-HDL, HbA1c, homeostasis model assessment of insulin resistance (HOMA-IR) and β-cell function (HOMA-β). We first modelled T2D-related traits as outcome variables in linear regression with age, sex BMI and obtained the residuals of each T2D-related trait respectively, then we calculated the Spearman correlation coefficients between the genera and the residuals of T2D-related traits. Results displaying a *p*<0.05 after Bonferroni adjustment were considered as significant.

## RESULTS

### Characteristics of the study participants

Table 1 and Table S1 summarize the baseline characteristics of the population by quartiles of erythrocyte GLA, LA and AA. The median levels (interquartile range) of baseline erythrocyte LA, GLA, AA and total n-6 PUFAs (% total fatty acids) were 9.84 (8.89-10.76), 0.037 (0.025-0.052), 11.51 (10.00-12.60) and 21.6 (19.6-23.1), respectively. Table S2 and S3 present baseline population characteristics of participants with and without follow-up information, 16S profiling respectively. Participants who dropped out had higher serum glucose levels, consumed fewer vegetables and energy, tended to be older, less educated and physically active, and were more likely to smoke.

**Table 1.**
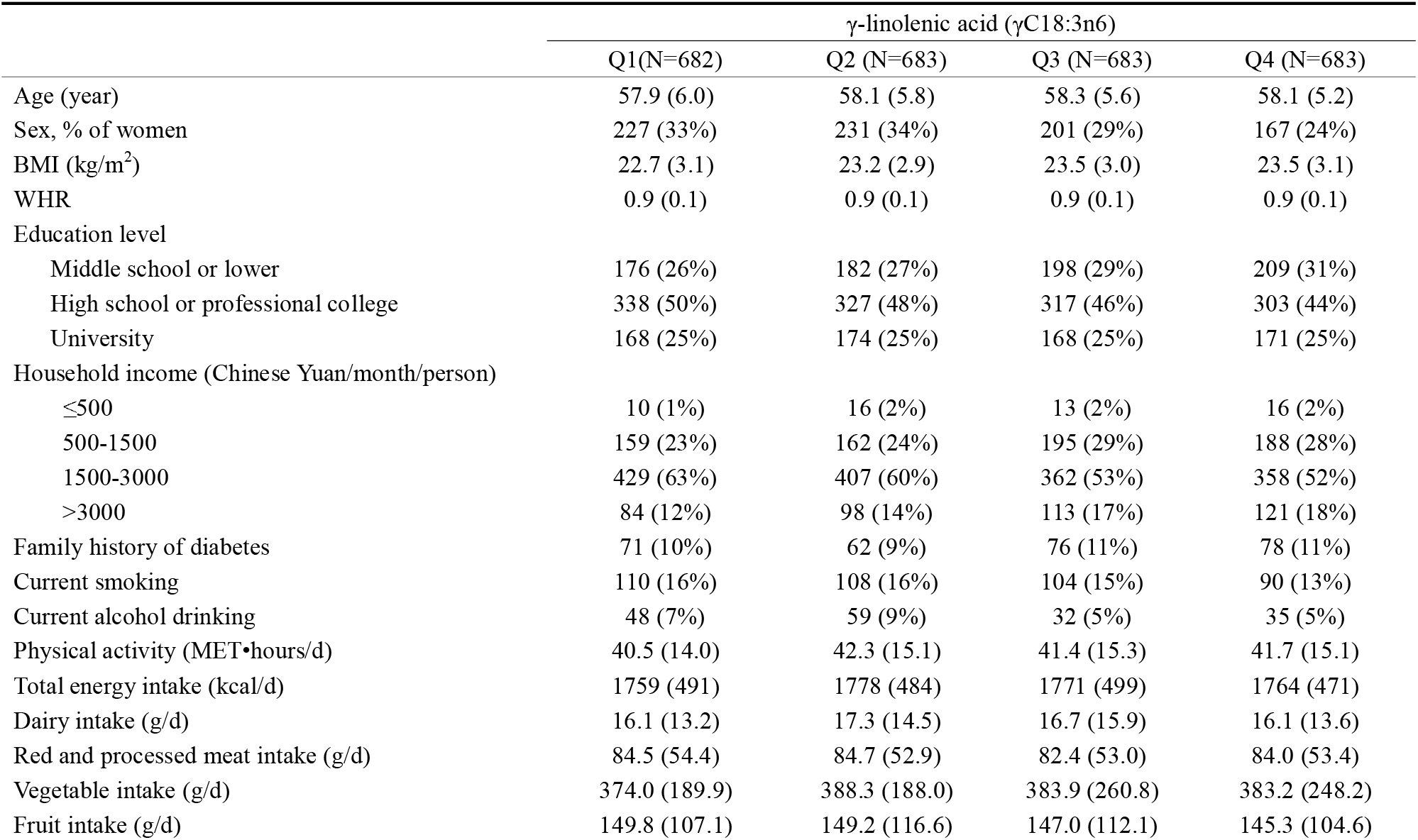
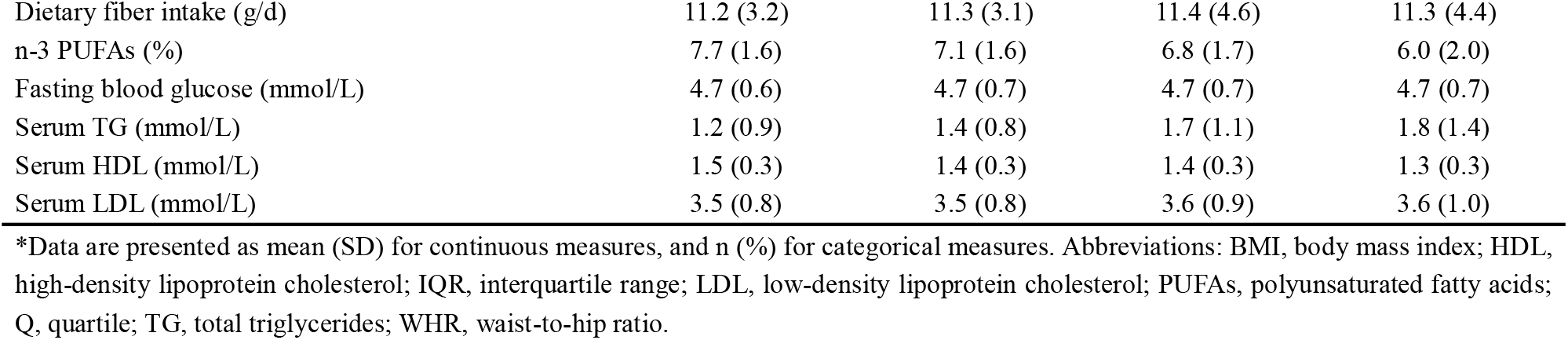
Baseline population characteristics by quartiles of erythrocyte γ-linolenic acid*

| | γ-linolenic acid (γC18:3n6) | | | |
| --- | --- | --- | --- | --- |
|  | Q1(N=682) | Q2 (N=683) | Q3 (N=683) | Q4 (N=683) |
| Age (year) | 57.9 (6.0) | 58.1 (5.8) | 58.3 (5.6) | 58.1 (5.2) |
| Sex, % of women | 227 (33%) | 231 (34%) | 201 (29%) | 167 (24%) |
| BMI (kg/m<sup>2</sup>) | 22.7 (3.1) | 23.2 (2.9) | 23.5 (3.0) | 23.5 (3.1) |
| WHR | 0.9 (0.1) | 0.9 (0.1) | 0.9 (0.1) | 0.9 (0.1) |
| Education level |  |  |  |  |
| Middle school or lower | 176 (26%) | 182 (27%) | 198 (29%) | 209 (31%) |
| High school or professional college | 338 (50%) | 327 (48%) | 317 (46%) | 303 (44%) |
| University | 168 (25%) | 174 (25%) | 168 (25%) | 171 (25%) |
| Household income (Chinese Yuan/month/person) |  |  |  |  |
| ≤500 | 10 (1%) | 16 (2%) | 13 (2%) | 16 (2%) |
| 500-1500 | 159 (23%) | 162 (24%) | 195 (29%) | 188 (28%) |
| 1500-3000 | 429 (63%) | 407 (60%) | 362 (53%) | 358 (52%) |
| >3000 | 84 (12%) | 98 (14%) | 113 (17%) | 121 (18%) |
| Family history of diabetes | 71 (10%) | 62 (9%) | 76 (11%) | 78 (11%) |
| Current smoking | 110 (16%) | 108 (16%) | 104 (15%) | 90 (13%) |
| Current alcohol drinking | 48 (7%) | 59 (9%) | 32 (5%) | 35 (5%) |
| Physical activity (MET•hours/d) | 40.5 (14.0) | 42.3 (15.1) | 41.4 (15.3) | 41.7 (15.1) |
| Total energy intake (kcal/d) | 1759 (491) | 1778 (484) | 1771 (499) | 1764 (471) |
| Dairy intake (g/d) | 16.1 (13.2) | 17.3 (14.5) | 16.7 (15.9) | 16.1 (13.6) |
| Red and processed meat intake (g/d) | 84.5 (54.4) | 84.7 (52.9) | 82.4 (53.0) | 84.0 (53.4) |
| Vegetable intake (g/d) | 374.0 (189.9) | 388.3 (188.0) | 383.9 (260.8) | 383.2 (248.2) |
| Fruit intake (g/d) | 149.8 (107.1) | 149.2 (116.6) | 147.0 (112.1) | 145.3 (104.6) |
| Dietary fiber intake (g/d) | 11.2 (3.2) | 11.3 (3.1) | 11.4 (4.6) | 11.3 (4.4) |
| n-3 PUFAs (%) | 7.7 (1.6) | 7.1 (1.6) | 6.8 (1.7) | 6.0 (2.0) |
| Fasting blood glucose (mmol/L) | 4.7 (0.6) | 4.7 (0.7) | 4.7 (0.7) | 4.7 (0.7) |
| Serum TG (mmol/L) | 1.2 (0.9) | 1.4 (0.8) | 1.7 (1.1) | 1.8 (1.4) |
| Serum HDL (mmol/L) | 1.5 (0.3) | 1.4 (0.3) | 1.4 (0.3) | 1.3 (0.3) |
| Serum LDL (mmol/L) | 3.5 (0.8) | 3.5 (0.8) | 3.6 (0.9) | 3.6 (1.0) |

\*Data are presented as mean (SD) for continuous measures, and n (%) for categorical measures. Abbreviations: BMI, body mass index; HDL, high-density lipoprotein cholesterol; IQR, interquartile range; LDL, low-density lipoprotein cholesterol; PUFAs, polyunsaturated fatty acids; Q, quartile; TG, total triglycerides; WHR, waist-to-hip ratio.

### Association of erythrocyte n-6 PUFAs with incident T2D

Higher levels of baseline GLA were associated with higher risk of T2D across the three statistical models (*p*-trend < 0.001 for all models, Table 2). For model 3, compared with the first quartile group (Q1), RRs (95% CIs) of T2D at Q2, Q3 and Q4 were 1.22 (0.85, 1.74), 1.43 (1.01, 2.03) and 1.72 (1.21, 2.44), respectively. No association was found for LA or AA. Similar results were obtained after using the simple or multiple imputation for the missing data in the sensitivity analysis (Table S4). Adjustment for additional dietary factors and disease histories showed similar results (Table S5). We did not observe significant interaction for n-6 PUFA biomarkers with age, sex, BMI or n-3 PUFAs on T2D risk (*p*_interaction_>0.05).

**Table 2.**
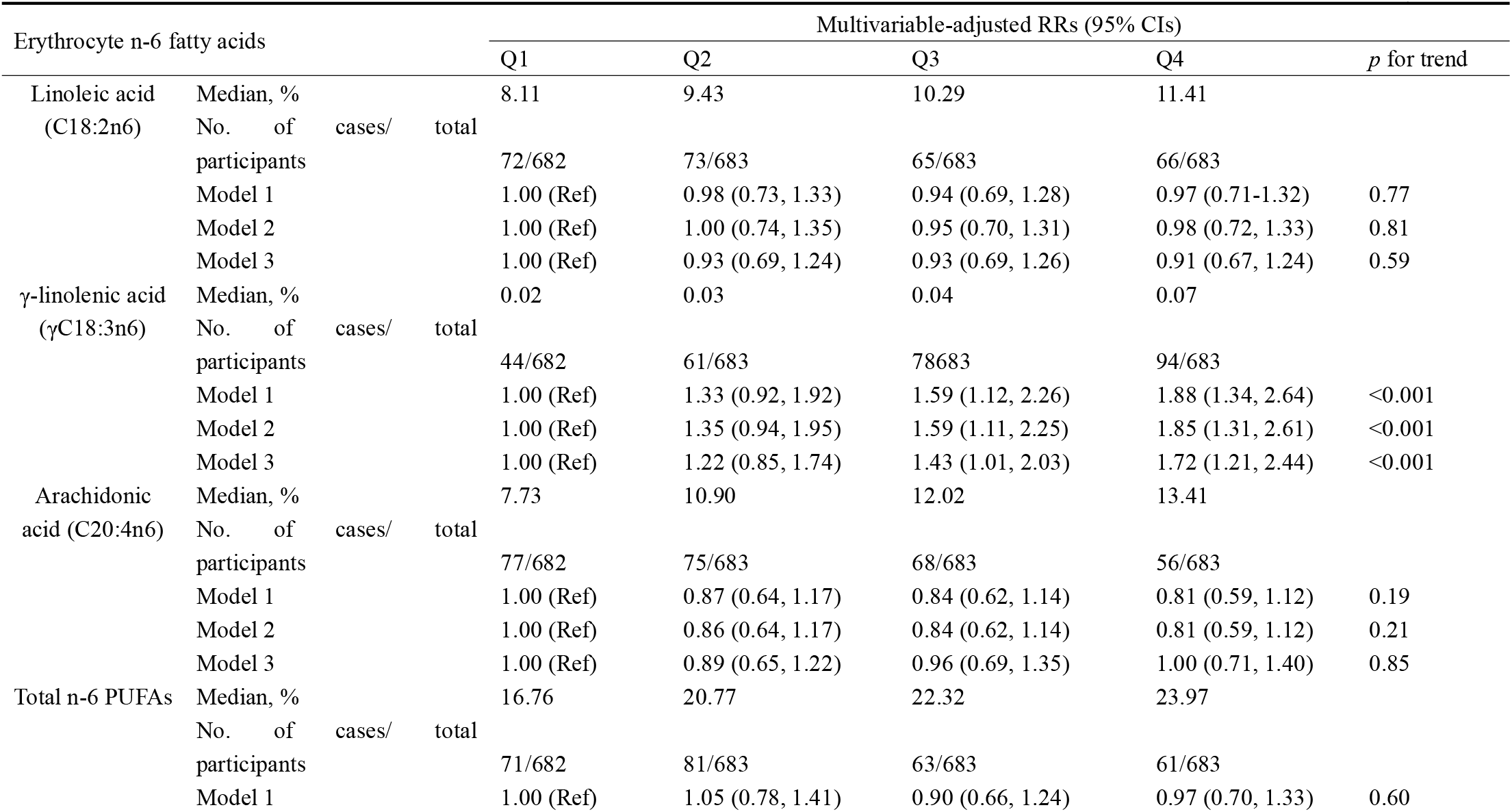
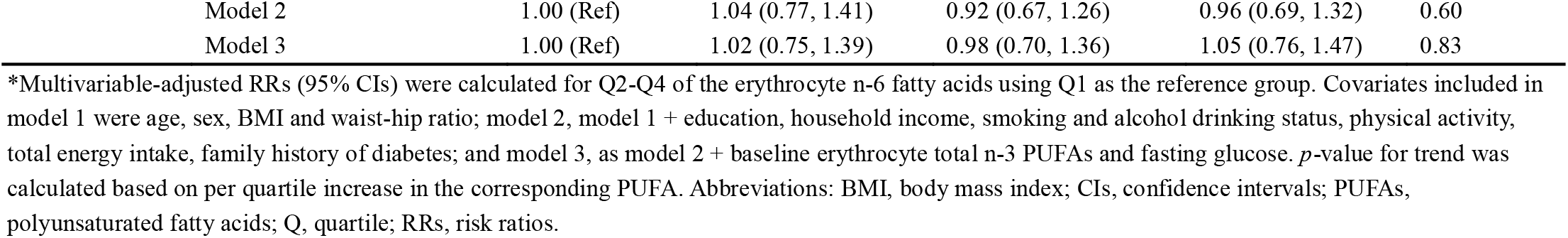
Association of erythrocyte n-6 fatty acids with incident type 2 diabetes*

| Erythrocyte n-6 fatty acids |  |  |  | Multivariable-adjusted RRs (95% CIs) |  |  |  | *p* for trend |
| --- | --- | --- | --- | --- | --- | --- | --- | --- |
|  |  |  |  | Q1 | Q2 | Q3 | Q4 |  |
| Linoleic acid<br>(C18:2n6) | Median, % |  |  | 8.11 | 9.43 | 10.29 | 11.41 |  |
|  | No. of cases/total participants |  |  | 72/682 | 73/683 | 65/683 | 66/683 |  |
|  | Model 1 |  |  | 1.00 (Ref) | 0.98 (0.73, 1.33) | 0.94 (0.69, 1.28) | 0.97 (0.71-1.32) | 0.77 |
|  | Model 2 |  |  | 1.00 (Ref) | 1.00 (0.74, 1.35) | 0.95 (0.70, 1.31) | 0.98 (0.72, 1.33) | 0.81 |
|  | Model 3 |  |  | 1.00 (Ref) | 0.93 (0.69, 1.24) | 0.93 (0.69, 1.26) | 0.91 (0.67, 1.24) | 0.59 |
| γ-linolenic acid<br>(γC18:3n6) | Median, % | | | 0.02 | 0.03 | 0.04 | 0.07 | |
|  | No. of cases/total participants |  |  | 44/682 | 61/683 | 78/683 | 94/683 |  |
|  | Model 1 |  |  | 1.00 (Ref) | 1.33 (0.92, 1.92) | 1.59 (1.12, 2.26) | 1.88 (1.34, 2.64) | <0.001 |
|  | Model 2 |  |  | 1.00 (Ref) | 1.35 (0.94, 1.95) | 1.59 (1.11, 2.25) | 1.85 (1.31, 2.61) | <0.001 |
|  | Model 3 |  |  | 1.00 (Ref) | 1.22 (0.85, 1.74) | 1.43 (1.01, 2.03) | 1.72 (1.21, 2.44) | <0.001 |
| Arachidonic acid (C20:4n6) | Median, % |  |  | 7.73 | 10.90 | 12.02 | 13.41 |  |
|  | No. of cases/total participants |  |  | 77/682 | 75/683 | 68/683 | 56/683 |  |
|  | Model 1 |  |  | 1.00 (Ref) | 0.87 (0.64, 1.17) | 0.84 (0.62, 1.14) | 0.81 (0.59, 1.12) | 0.19 |
|  | Model 2 |  |  | 1.00 (Ref) | 0.86 (0.64, 1.17) | 0.84 (0.62, 1.14) | 0.81 (0.59, 1.12) | 0.21 |
|  | Model 3 |  |  | 1.00 (Ref) | 0.89 (0.65, 1.22) | 0.96 (0.69, 1.35) | 1.00 (0.71, 1.40) | 0.85 |
| Total n-6 PUFAs | Median, % |  |  | 16.76 | 20.77 | 22.32 | 23.97 |  |
|  | No. of cases/total participants |  |  | 71/682 | 81/683 | 63/683 | 61/683 |  |
|  | Model 1 |  |  | 1.00 (Ref) | 1.05 (0.78, 1.41) | 0.90 (0.66, 1.24) | 0.97 (0.70, 1.33) | 0.60 |

|  |  |  |  |  |  |
| --- | --- | --- | --- | --- | --- |
| Model 2 | 1.00 (Ref) | 1.04 (0.77, 1.41) | 0.92 (0.67, 1.26) | 0.96 (0.69, 1.32) | 0.60 |
| Model 3 | 1.00 (Ref) | 1.02 (0.75, 1.39) | 0.98 (0.70, 1.36) | 1.05 (0.76, 1.47) | 0.83 |

\*Multivariable-adjusted RRs (95% CIs) were calculated for Q2-Q4 of the erythrocyte n-6 fatty acids using Q1 as the reference group. Covariates included in model 1 were age, sex, BMI and waist-hip ratio; model 2, model 1 + education, household income, smoking and alcohol drinking status, physical activity, total energy intake, family history of diabetes; and model 3, as model 2 + baseline erythrocyte total n-3 PUFAs and fasting glucose. *p*-value for trend was calculated based on per quartile increase in the corresponding PUFA. Abbreviations: BMI, body mass index; CIs, confidence intervals; PUFAs, polyunsaturated fatty acids; Q, quartile; RRs, risk ratios.

### The prospective association of erythrocyte n-6 PUFA biomarkers with gut microbiota diversity

Lower GLA was associated with higher gut microbiota α-diversity (Q4 *vs*. Q1: *p*=0.021 for observed OTUs, 0.028 for Shannon’s diversity index, 0.023 for Chao index, respectively) (Figure 1). However, baseline LA, AA or total n-6 PUFAs were not associated with the microbial α-diversity (Table S6). These results remained similar after further adjusting for dietary fiber intake (Table S7). Among 1,591 individuals included in the microbial association analysis, we also observed positive association between GLA and T2D (Table S8). α-diversity acted as a potential partial mediator of the association between GLA and T2D (Table 3). For β-diversity, GLA and AA significantly (*p*<0.01) contributed to dissimilarities in gut microbiota composition at genus level (Table S9). Specifically, for GLA, the gut microbiota composition at genus level was different across quartile groups of GLA concentrations (Table S10), with *p* = 0.006 for comparison between Q1 and Q4, for example, as visualized in Figure 2.

**Table 3.** Summary statistics of the mediation analysis for α-diversity indicators*

|  | **n** | **ACME** | ***p*** | **ADE** | ***p*** | **%** |
| --- | --- | --- | --- | --- | --- | --- |
| Observed OTUs | 1541 | 0.0011 | 0.006 | 0.14 | 0.01 | 7.9 |
| Chao index | 1541 | 0.00097 | 0.011 | 0.012 | 0.0093 | 7.1 |
| Shannon's diversity index | 1541 | 0.00098 | 0.005 | 0.012 | 0.01 | 7.1 |

\* Models were built to test whether the association between γ-linolenic acid and type 2 diabetes was mediated by gut microbial α-diversity indices. The direct and indirect effect are calculated using the quasi-Bayesian Monte Carlo method with 2,000 simulations (R *{mediation}*). Covariates included age, sex, BMI and waist-hip ratio, education, household income, smoking and alcohol drinking status, physical activity, total energy intake, family history of diabetes, baseline erythrocyte total n-3 polyunsaturated fatty acids and fasting glucose. Abbreviations: BMI, body mass index.

**Figure 1.**
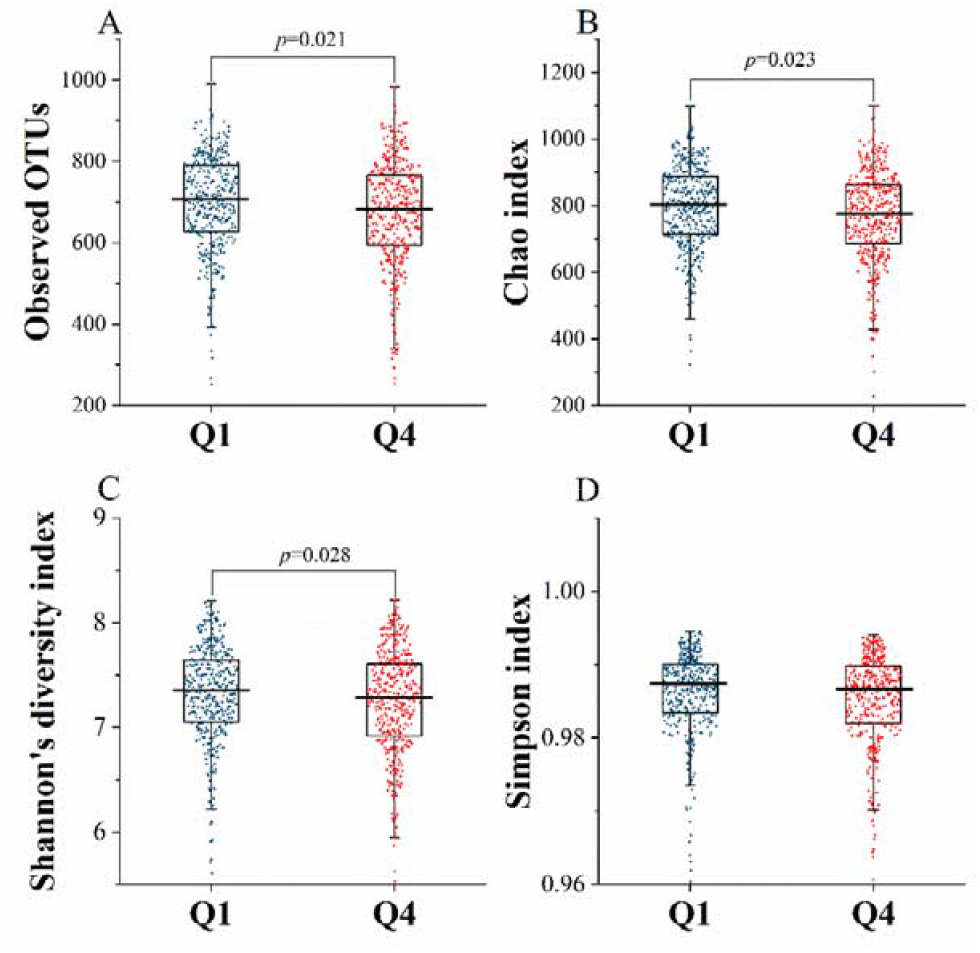
Gut microbial community richness and diversity between quartile 1 and quartile 4 of erythrocyte γ-linolenic acid. The box plots feature the median (centerline), upper and lower quartiles (box limits), 1.5× the interquartile range (whiskers). Gut microbial community richness was represented by Observed OTUs (A) and Chao index (B), and community diversity was represented by Shannon’s diversity index (C) and Simpson index (D). *p* values were calculated for Q4 (n=397) of the erythrocyte n-6 fatty acids using Q1 (n=398) as the reference group using a linear mixed model: sequencing batch as random effect; sequencing depth, Bristol scale, age, sex, BMI, waist-hip ratio, education, household income, smoking and alcohol drinking status, physical activity, total energy intake, family history of diabetes, baseline erythrocyte total n-3 PUFAs and fasting glucose as fixed effects. Abbreviations: BMI, body mass index, Q, quartile.

**Figure 2.**
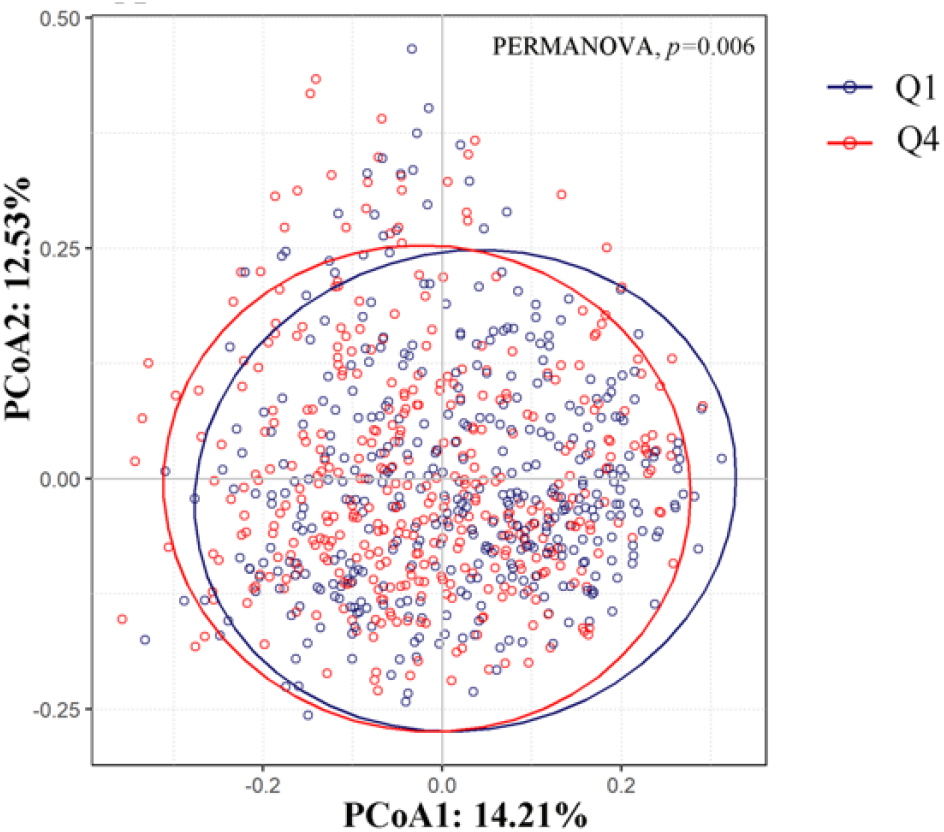
Dissimilarities in gut microbiota composition (β-diversity) between quartile 1 and quartile 4 of erythrocyte γ-linolenic acid. Dissimilarities in gut microbiota composition between quartile 1 and quartile 4 of erythrocyte γ-linolenic acid (n=795) represented by unconstrained PCoA with the Bray-Curtis dissimilarity index calculated on scaled relative abundances at genus level. Baseline erythrocyte γ-linolenic acid level explained 0.3% of the dissimilarities in gut microbiota composition (PERMANOVA, *p*= 0.002). Abbreviation: Q, quartile.

### The cross-sectional association of gut microbiota diversity with T2D

T2D patients had a lower α-diversity represented by observed OTUs, Shannon’s diversity index, Simpson index and Chao index compared with patients without T2D (Table S11). We also found that the overall β-diversity was different between participants with and without T2D (Table S12).

### Taxonomic profiles of gut microbial community in different levels of GLA

We then focused on exploring gut genera potentially underlying the association between GLA and incident T2D. At the genus level, the gut microbiota of participants within Q1 and Q4 of GLA was both dominated by *Bacteroides*, while the relative abundance of *Bacteroides* was significantly higher in Q4 group (*p*=0.016). We also identified *Rothia, [Eubacterium], Coprococcus* enriched in participants with high GLA levels (Q4) using LDA method (Figure 3). We found several overlapping taxonomic biomarkers for non-T2D status and low GLA status, including *Butyrivibrio, Blautia, Oscillospira, Odoribacter, S24-7 other, Rikenellaceae other*, and *Clostridiales other* (Figure 3). In the cross-sectional analysis relating taxonomic factors to metabolic traits, the genus *[Eubacterium], S24-7 other, Blautia, Oscillospira, Odoribacter, Rikenellaceae other, Coriobacteriaceae Other, Faecalibacterium* and *Christensenellaceae Other* were negatively correlated with total triglycerides (Bonferroni corrected *p*<0.05) (Figure 4). No correlation was observed for other metabolic trait-microbiota pairs (Table S13).

**Figure 3.**
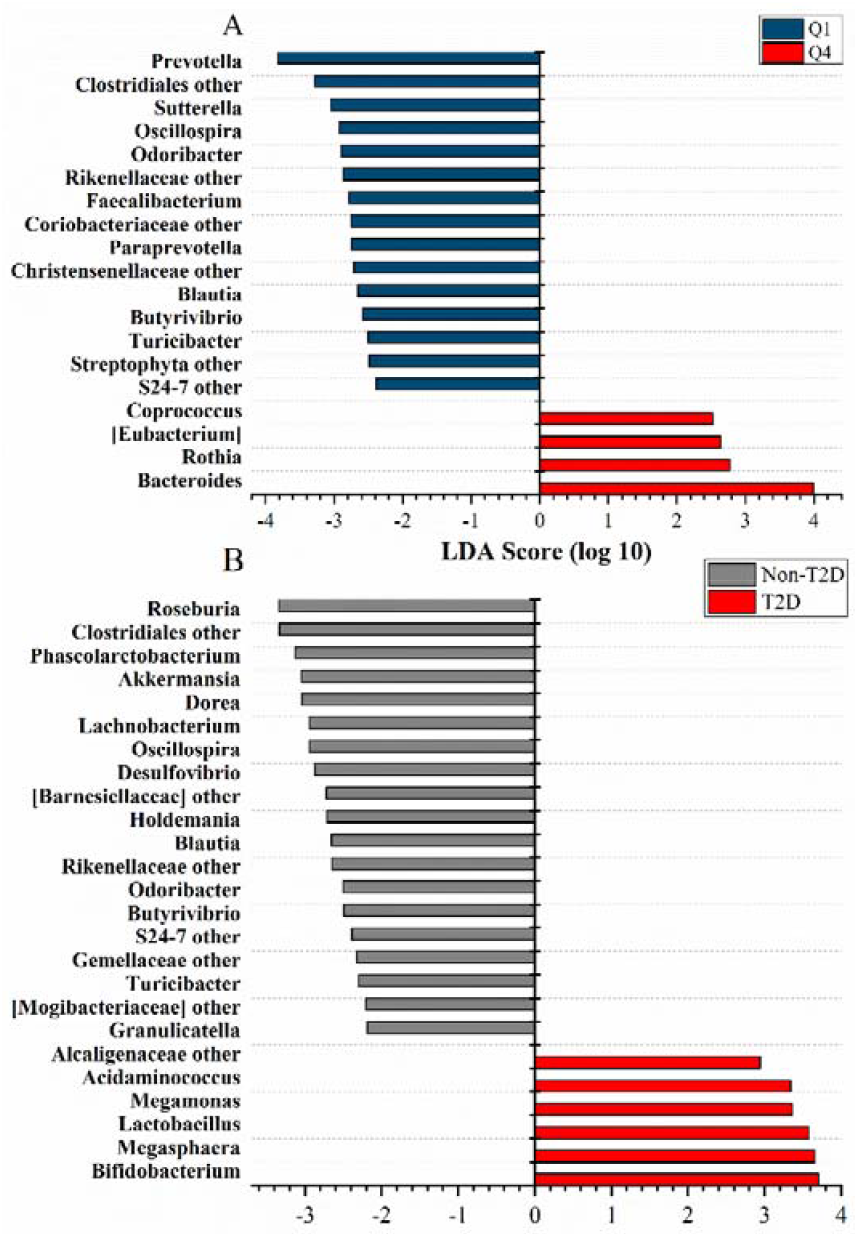
Gut microbial biomarkers at genus level by γ-linolenic acid and type 2 diabetes. Gut microbial taxonomic biomarkers identified by the linear discriminant analysis (LDA) at quartile 1 and 4 of erythrocyte γ-linolenic acid (A), and by type 2 diabetes status (B). Color indicates the group in which a differentially abundant taxon is enriched ((A) dark blue: quartile 1 of GLA; red: quartile 4 of GLA; (B) gray: participants without T2D; red: participants with T2D). Abbreviation: Q, quartile; T2D, type 2 diabetes; Non-T2D, participants without type 2 diabetes.

**Figure 4.**
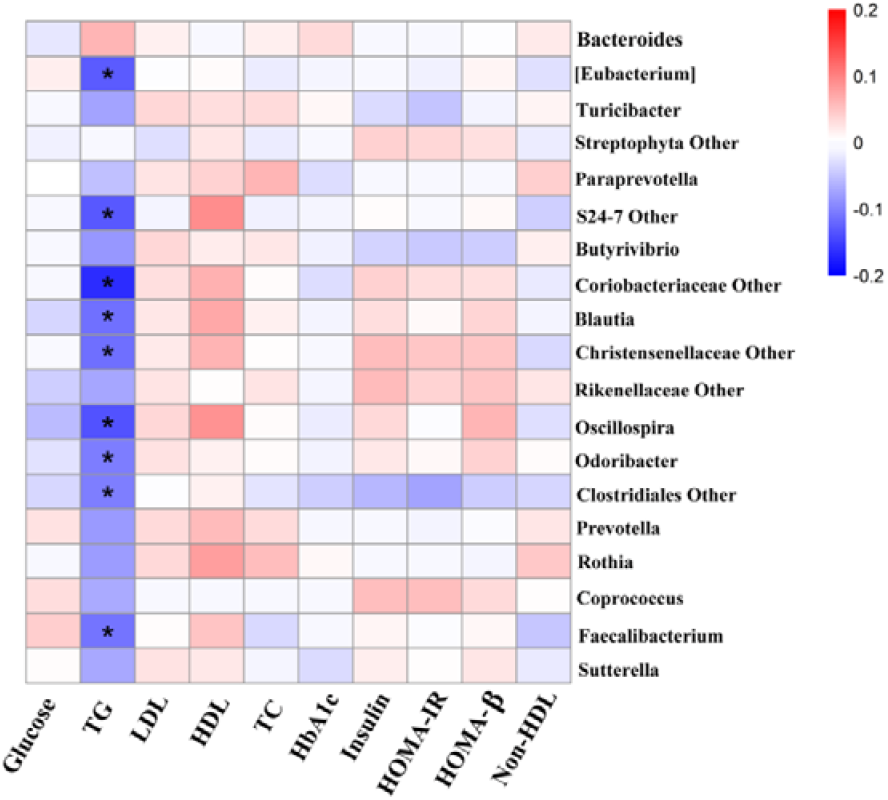
Heatmap of the Spearman correlation coefficients between γ-linolenic acid-related microbes and T2D-related traits. The intensity of the colors represents the degree of association between γ-linolenic acid-related microbes and ten T2D-related traits as measured by the Spearman’s correlations. All significant correlations are marked with asterisk (Bonferroni corrected *p*<0.05). Abbreviation: TG, total triglycerides; TC, total cholesterol; LDL, low-density lipoprotein cholesterol; HDL, high-density lipoprotein cholesterol; HOMA-IR, homeostasis model assessment of insulin resistance; HOMA-β, homeostasis model assessment of β-cell function; T2D, type 2 diabetes.

## DISCUSSION

In the present community-based prospective cohort of a Chinese population, we found that baseline erythrocyte GLA levels were positively associated with incident T2D, independent of BMI and other potential confounders. Proportions of LA, AA or total n-6 PUFAs were not associated with T2D incidence. Our further investigation integrating gut microbiota data revealed that GLA may be associated with T2D risk through a mechanism which varies diversity and composition of gut microbiota. Our findings suggest that n-6 PUFAs and gut microbiota co-vary in the development of T2D risk, highlighting the presence of a novel mechanism of how fatty acids or gut microbiota influence T2D risk.

Our findings suggest that high circulating GLA might be a risk factor of T2D, which are consistent with several prospective studies in western countries.^23 24^ The null finding for AA in our present study is also consistent with those from the previous studies.^5 23 25^ The findings for LA were inconsistent between ours and previous ones.^5 23^ Circulating LA reflects habitual consumption of n-6 PUFA, typically from plant-derived oils. The inconsistency may reflect that European and Chinese populations use plant oils differently with a combination of different foods. For instance, Chinese adults may consume plant oils with meat by which positive and negative effects of those would lead to the null finding for LA. By contrast, an American or European population may use plant oils with vegetables or in a healthy diet pattern. Such dietary confounding might influence GLA finding little, as GLA is a product of tightly-regulated desaturation of LA, and result in the consistent findings between studies.

Given the putative relationship between gut microbiome and T2D, we hypothesized and confirmed that gut microbiome diversity may play a role linking n-6 PUFAs and T2D and that n-6 PUFAs may prospectively affect gut microbiota composition. Our findings are in line with a randomized controlled trial in which α-diversity, *Blautia* was reduced, while *Bacteroides* were increased after higher-fat diet intervention (rich in n-6 PUFAs owing to exclusive use of soybean oil).^26^ Although we did not observe higher relative abundance of *Bacteroides* in T2D patients in our study, *Bacteroides* has been found to be more abundant in T2D in another Chinese population.^9^

We found that *Butyrivibrio, Blautia, Oscillospira*, and *Odoribacter*, genera known to contain a series of butyrate-producing bacteria, are all enriched in participants within Q1 of GLA and participants without T2D. *Butyrivibrio fibrisolvens* was reported to have the ability to utilize fructose polymers and it showed anticarcinogenic effect in mice.^27 28^ *Blautia* was reported to have a lower relative abundance in T2D patients and have inverse association with visceral fat.^29 30^ *Oscillospira* was positively associated with leanness and was reduced in inflammatory diseases.^31^ In addition, evidence suggested that some species of *Oscillospira* were able to utilize glucuronate, a common animal-derived sugar.^32^ *Odoribacter splanchnicus* was considered as a potential probiotics candidate for obesity prevention as a higher abundance in obesity-depleted group.^33^ Moreover, in our study, *Blautia, Oscillospira* and *Odoribacter* were inversely associated with circulating triglycerides, which was found to be positively associated with T2D and higher level of GLA.^34 35^ Thus, our study suggests that lower level of GLA may be correlated with factors increasing the abundance of these bacteria and preventing the development of T2D.

The more detailed mechanism behind these associations remains unclear but may be related to chronic inflammation. Accumulating evidence supports the hypothesis that chronic low-grade inflammation is a risk factor for the development of T2D,^36^ and that n-6 PUFA may play a vital role in the regulation of inflammation.^37^ GLA is produced from LA by the enzyme delta-6-desaturase, and is further metabolized to dihomo-γ-linolenic acid (DGLA; 20:3n6) which undergoes oxidative metabolism by cyclooxygenases and lipoxygenases to produce anti-inflammatory eicosanoids such as prostaglandins and leukotrienes.^38^ Moreover, imbalance of the gut microbiota and some specific microbes have been considered as contributors for inflammation by gut microbiota-derived lipopolysaccharide(LPS)^39 40^. The LPS stimulates Toll-like receptors (TLRs) to influence innate immunity via its interaction with LPS receptors (i.e., CD14/TLR 4complex).^41^ It can be proposed that LPS level is regulated by fat and fatty acids, and associated with insulin resistance and T2D.^40 42 43^ Moreover, the enrichment of inflammation-related bacteria (i.e., *Blautia*^44 45^) in participants with lower levels of GLA supports an important role of inflammation in the relationship between GLA, gut microbiota and T2D.

Strengths of the present study include the prospective cohort study design, which is rarely conducted in prior research on fatty acid-gut microbiota associations, and the collection and adjustment for a variety of information including socio-demographic, lifestyle and dietary factors, anthropometric parameters and circulating blood biomarkers. Several limitations merit attention. First, although we have adjusted for several potential confounders, due to the observational nature of the present study, we are not able to fully exclude the influence of residual confounding. Second, erythrocyte n-6 PUFAs were measured at baseline only and may not represent long-term status. Third, fecal samples were only collected during follow-up but not at baseline. Repeated measure of both circulating n-6 PUFAs and gut microbiota or an intervention of n-6 PUFAs would enable us to characterize bidirectional effects of n-6 PUFA intakes on gut microbiota and of gut microbiota on circulating n-6 PUFAs. Nevertheless, our study fills the gap of the prospective associations between n-6 PUFA biomarkers and gut microbiota.

In conclusion, our study suggests that erythrocyte GLA biomarker is positively associated with incident T2D in the Chinese population. Gut microbiota may play an important role to explain the finding. The identification of GLA-related gut microbiota features may serve as potential intervention targets for further investigation. The detailed mechanism behind the association of GLA with T2D risk and gut microbiota needs further clarification in the future.

## Data Availability

The raw data for 16â∈%S rRNA gene sequences are available in the CNSA (https://db.cngb.org/cnsa/) of CNGBdb at accession number CNP0000829.

## Acknowledgments

We thank all the participants and staff of the GNHS study. We thank Westlake University Supercomputer Center for computational resource and related assistance.

## Contributors

JSZ, YMC conceived the study concept and design. GDC, FFZ, HLD, JW, CX collected the data. ZM, JSL, YM did the data analysis. JSZ, ZM, JSL, YM wrote the first draft of the article. ZJ, MS, WG, YF, FI, YMC contributed to the discussion and critical revision of the manuscript. All authors read, revised and approved the final draft.

## Patient consent

Obtained

## Ethics approval

This study was approved by the Ethics Committee of the School of Public Health at Sun Yat-sen University (2018048) and Ethics Committee of Westlake University (20190114ZJS0003).

